# Understanding Psychiatric Illness Through Natural Language Processing (UNDERPIN): Rationale, Design, and Methodology

**DOI:** 10.1101/2021.12.05.21267037

**Authors:** Taishiro Kishimoto, Hironobu Nakamura, Yoshinobu Kano, Yoko Eguchi, Momoko Kitazawa, Kuo-ching Liang, Koki Kudo, Ayako Sento, Akihiro Takamiya, Toshiro Horigome, Toshihiko Yamasaki, Yuki Sunami, Toshiaki Kikuchi, Kazuki Nakajima, Masayuki Tomita, Shogyoku Bun, Yuki Momota, Kyosuke Sawada, Junichi Murakami, Hidehiko Takahashi, Masaru Mimura, on behalf of the UNDERPIN Collaborators

## Abstract

**Introduction:** Psychiatric disorders are diagnosed according to diagnostic criteria such as the DSM-5 and ICD-11. Basically, psychiatrists extract symptoms and make a diagnosis by conversing with patients. However, such processes often lack objectivity. In contrast, specific linguistic features can be observed in some psychiatric disorders, such as a loosening of associations in schizophrenia. The purposes of the present study are to quantify the language features of psychiatric disorders and neurocognitive disorders using natural language processing and to identify features that differentiate disorders from one another and from healthy subjects.

**Methods:** This study will have a multi-center prospective design. Major depressive disorder, bipolar disorder, schizophrenia, anxiety disorder including obsessive compulsive disorder and, major and minor neurocognitive disorders, as well as healthy subjects will be recruited. A psychiatrist or psychologist will conduct 30-to-60-min interviews with each participant and these interviews will be recorded using a microphone headset. In addition, the severity of disorders will be assessed using clinical rating scales. Data will be collected from each participant at least twice during the study period and up to a maximum of five times.

**Discussion:** The overall goal of this proposed study, the Understanding Psychiatric Illness Through Natural Language Processing (UNDERPIN), is to develop objective and easy-to-use biomarkers for diagnosing and assessing the severity of each psychiatric disorder using natural language processing. As of August 2021, we have collected a total of >900 datasets from >350 participants. To the best of our knowledge, this data sample is one of the largest in this field.

**Trial registration:** UMIN000032141, University Hospital Medical Information Network (UMIN).

## 1. Introduction

Psychiatric disorders have a large impact on humans and can reduce quality of life (QOL) because of a high incidence and long duration of illness. According to global burden of disease surveys conducted by the World Health Organization (WHO) and other organizations, psychiatric disorders, including depression, bipolar disorder, anxiety disorder, schizophrenia, and drug addiction, are the leading medical disorders in terms of years lived with disability (YLDs), accounting for 18.7% of the global YLD in 2019 [1].

Psychiatrists diagnose psychiatric disorders by conducting one-to-one conversations with each patient. Historically, heuristic studies such as linguistics and psychopathology have vigorously studied the thought and language of psychiatric disorders. Several linguistic features are known, such as a loosening of associations in schizophrenia, flight of ideas in mania, psychomotor inhibition in depression, and circumstantial and word recall disorder in dementia. Although there is a common understanding of these linguistic features among psychiatrists, the diagnosis and evaluation of the degree to which a patient deviates from the range of what is considered healthy depends heavily on each psychiatrist’s sensitivity and experience. One of the reasons why evaluations remain subjective is that there are no means of quantifying psychiatric diseases to date. These subjective judgments can lead to various problems, such as diagnostic disagreements among psychiatrists, unclear criteria for initiating treatment, and difficulty providing a standardized education of resident doctors. Although the characteristics of the disease appear in each patient’s words, it is sometimes difficult to diagnose atypical cases or cases with very severe symptoms, as it can be difficult to determine what symptoms are causing the difficulty in coherent speech. This research aims to deepen our understanding of the characteristics of psychiatric disorders and to quantify them using natural language processing and with machine learning so that in the future, the features of psychiatric disorders can be identified and quantified. Such technology could eventually be used for early diagnosis and prevention.

## 2. Methods

### 2.1. Participants

This study is a multicenter, prospective, observational study. Subject recruitment was started in 2018 and is ongoing. Participants are being recruited at seven hospitals and three outpatient clinics in Japan. Patient recruitment is being conducted at the following locations and hospitals: Tokyo (Keio University Hospital, Tsurugaoka Garden Hospital, Oizumi Hospital, Komagino Hospital, Oizumi Mental Clinic, Asakadai Mental Clinic), Kanagawa (Nagatsuta Ikoinomori Clinic), Shiga (Biwako Hospital), Yamagata (Sato Hospital), and Fukushima (Asaka Hospital). Participants are inpatients or outpatients aged ≥20 years who meet the diagnostic criteria for depression, bipolar disorder, schizophrenia, anxiety disorder (including obsessive-compulsive disorder), mild cognitive impairment, or dementia according to the DSM-5 or ICD-11. Healthy volunteers will consist of healthy individuals with no history of psychiatric disorders who have volunteered to participate in the study after reading the research group’s website and printed recruitment advertisements, and who are at least 20 years old at the time of consent. Researchers will obtain written informed consent from all the participants.

### 2.2. Data collection

The following data will be collected in this study.

#### Demographic characteristics

Sex, age, diagnosis, diagnosis subtype, duration of illness, prescription, and complications will be collected from the medical records. If some information is missing, the patients will be asked to provide the information.

#### Conversation data

A 60-minute conversation with a study psychiatrist or psychologist will be conducted. The content of the conversation will be similar to a typical 30-to-60-minute conversation conducted in psychiatric clinical practice. Questions will be asked about the course of the illness, the patient’s recent condition, daily life, sleep patterns, interests and hobbies, and comments on news, movies, stories, pictures, etc. No strict structuring will be performed. The interviewer and patients will be asked to wear microphone headsets, and the conversation will be audio-recorded.

#### Clinical assessment

The severity of the illness will be assessed using the rating scales described in the table (Table1). In addition, the treating psychiatrists will be asked to evaluate each patient’s recent disease severity using the Clinical General Impression Scale-Severity (CGI-S). During the follow-up interview, a structured psychiatric diagnostic interview (SCID) will be conducted to the greatest degree feasible to confirm the diagnoses.

**Table1.** Data Collection and Visit Schedule

|  |  |  | 1 | 2 | 3 | 4 | 5 |
| --- | --- | --- | --- | --- | --- | --- | --- |
|  |  |  | Assessments must be done in<br>Visit 1 and Visit 2 |  | Assessments will be done<br>as long as possible |  |  |
| A) Demographic characteristics | All participants | Demographic characteristics | ✓ |  |  |  |  |
|  |  | Information on treatment history | ✓ | ✓ | ✓ | ✓ | ✓ |
|  | Only patients | SCID | ✓ (Once during research period) |  |  |  |  |
| B) Conversation with a psychiatrist<br>or psychologist | All participants | Conversation similar to a daily<br>consultation | ✓ | ✓ | ✓ | ✓ | ✓ |
| C) Severity assessment using clinical<br>rating scales | C-1) Monopolar depression<br>disorder<br>or Bipolar disorder<br>or Anxiety disorder | HAM-D<br>MADRS<br>YMRS<br>HAM-A<br>STAI<br>Satisfaction With Life Scale<br>(SWLS)<br>Cantril's Ladder of life scale<br>The Flourishing Scale (FS-J)<br>Subjective Well-being Inventory | ✓ | ✓ | ✓ | ✓ | ✓ |
|  | C-2) Schizophrenia | PANSS<br>Satisfaction With Life Scale<br>(SWLS)<br>Cantril's Ladder of life scale<br>The Flourishing Scale (FS-J)<br>Subjective Well-being Inventory | ✓ | ✓ | ✓ | ✓ | ✓ |
|  | C-3) Neurocognitive<br>disorder | MMSE<br>CDR<br>MoCA-J<br>WMS-R LM<br>Satisfaction With Life Scale<br>(SWLS)<br>Cantril's Ladder of life scale<br>The Flourishing Scale(FS-J)<br>Philadelphia Geriatric Center Morale<br>Scale<br>Subjective Well-being Inventory | ✓ | ✓ | ✓ | ✓ | ✓ |
|  | For healthy volunteers | *M.I.N.I.<br>*MMSE<br>(*: Performed before all of the other tests) | ✓ |  |  |  |  |
|  |  | All of C-1), C-2), C-3) | ✓ | ✓ | ✓ | ✓ | ✓ |

Healthy volunteers will be screened using the Mini-International Neuropsychiatric Interview (M.I.N.I.) and the Mini-Mental State Examination (MMSE) and will be excluded if they have a history of psychiatric disorders or cognitive impairment. For the M.I.N.I., if any diagnosis is found, and for the MMSE, if the score is below 27, the participant will not be allowed to participate in the study as a healthy subject. After screening, the same assessments as those described above for the patients will also be conducted for the healthy subjects.

In this study, not only the collection of linguistic information associated with each disease, but also the relationship between illness severity and linguistic information will be important. We therefore will follow the participants two to five times during the study period in an attempt to interview the subjects at times when the severities of their symptoms differ (i.e., severe, moderate, mild, and during remittance). An interval of at least one month between the two assessments will be required. For the follow-up data collection, the same procedures described above will be conducted.

SCID: Structural Clinical Interview for DSM-5, HAM-D: Hamilton Depression Rating Scale, MADRS: Montgomery-Asberg Depression Rating Scale, YMRS: Young Mania Rating Scale, STAI: State-Trait Anxiety Inventory, PANSS: Positive And Negative Syndrome Scale, MMSE: Mini-Mental State Examination, MoCA-J: Montreal Cognitive Assessment-Japanese version, WMS-R LM: Wechsler Memory Scale-Revised Logical Memory, CDR: Clinical Dementia Rating, M.I.N.I.: Mini-International Neuropsychiatric Interview

### 2.3. Data processing and annotation

The acquired voice data will be converted into text information using speech recognition technology developed by the research group and will be compensated manually; annotations, such as pauses, fillers, nods, interjections, repetitions, co-supplementation, lexical responses, incomplete sentences, misstatements, etc., will also be made. At the same time, the following phonic information will be extracted from the recorded audio data to utilize the physical properties of the audio data in the machine learning model: fundamental frequency (F0); first, second, and third formant frequencies (F1, F2, F3); cepstral peak prominence (CPP); and mel-frequency cepstrum coefficients (MFCC). Software such as Praat [2]and openSMILE [3] will be used to extract such phonic information. The annotated text data thus obtained will then be replaced by various variables using natural language processing, such as morphological and syntactic analyses using MeCab [4, 5] and JUMAN [6, 7]. For instance, these analyses will include the frequency of the occurrence of each part of speech, vocabulary (negative and positive words), syntactic complexity, length of the utterance, frequency of occurrence of person, use of pronouns and distance to the referent, use of case structure (“te, ni, wo, ha”), and so on.

### 2.4. Statistical analysis and machine learning

#### Between group comparisons

Linguistic features considered to be characteristic of specific psychiatric diseases will be selected and statistically compared between groups (e.g., patients vs. healthy subjects, patients in different disease groups). This step is necessary to verify whether each variable has pathological validity before using it for machine learning.

#### Machine learning

The machine learning models in this study will be trained to perform the following tasks: 1) to predict whether a subject has or does not have psychiatric disorders such as depression, bipolar disorder, anxiety disorder, schizophrenia, or dementia; 2) to predict the severity of a subject’s state based on the results of rating scales such as the Hamilton Rating Scale for Depression (HAM-D), the Young Mania Rating Scale (YMRS) for bipolar disorder, the Positive And Negative Syndrome Scale (PANSS) for schizophrenia, and MMSE for dementia; 3) to predict the improvement or deterioration of a subject’s disorder with respect to a previously recorded state if the subject has undergone a prior assessment; and 4) to predict the scores of individual items in clinical rating scales that are indicators of different aspects of a subject’s disorder, such as positive symptoms, negative symptoms, and disorganized symptoms for schizophrenia. The data used to train these machine learning models may include voice features extracted from voice data obtained from subjects, conversational and linguistic information from transcribed text data, and annotations assigned to the data described above.

Next, we will need to perform feature engineering and to design the architecture of the machine learning. Data substituted into various variables using natural language processing will be used to extract disease features using machine learning such as decision trees, support vector machines (SVM), and deep learning. The importance of the features will then be verified using cross-validation and other methods. In addition to the accuracy of disease identification, we will simultaneously verify the possibility of estimating disease severity using support vector regression and other methods. We will then evaluate the results using the mean absolute error, coefficient of determination, and correlation coefficients. We also plan to verify whether the features obtained in this manner can not only distinguish between a normal and disease state or among different diseases, but also whether they can distinguish changes in disease severity over time.

### 2.5. Sample size

The results of a pilot study conducted on 30 patients prior to the main study showed that the combined natural language processing and machine learning had an accuracy of 72% to 80% for disease identification (based on a leave-one-out cross validation). Using 18 data points from a typical and ideally collected pilot study with subjects labeled as depression, dementia, and normal, we constructed a learning curve for classification and estimated the number of samples needed to achieve at least a 90% prediction accuracy in this study; we calculated that a minimum of 50 samples would be needed. In addition, considering the variability of the data over a wide range of age groups, severity of illness, and regional differences, it was estimated that about 10 times the number of samples would be necessary. Another means of approximating sample sizes for regression analysis is that the number of cases should be approximately 10 times the number of the independent variables used to obtain statistical confidence [8]. In the pilot study, we used 62-dimensional features for speech information and linguistic information, such as parts of speech and clauses. In the present study, we plan to use a total of about 100 independent variables for the analysis; based on the approximations, we estimate that the desired sample size is about 1,000. It should also be noted that the actual needed number of samples is likely to be lower because we will also be employing dimension reduction and feature selection techniques to reduce the number of independent variables in the model.

## 3. Discussion

The UNDERPIN study is unique in its purpose and the novelty of its method, such as the use of natural language processing, and in the size of data sample. In addition, it has a broad and evolving perspective that is expected to shed light on traditional psychiatry by applying the new analytical method of machine learning to language, a measure that has long been used as a cue in psychiatric practice.

Psychiatrists have long relied on language and superficial behavioral data to diagnose psychiatric disorders. Although many empirical studies have explored biomarkers of mental illness [9, 10], no robust biomarkers have been identified that are clinically practicable.

This study will focus on the linguistic features of psychiatric disorders, which have been explored using traditional disciplines such as linguistics [11, 12] and psychopathology. By introducing the method of natural language processing, we will be able to bring an empirical and mathematical perspective to humanistic examinations. Boer et al. identified features that distinguish patients with schizophrenia and healthy subjects based on speech data; using the calculated features, they then predicted the integrity of the white matter language tracts in two groups [13]. Although such studies remain rare, they may bring clinical practice and research on psychiatric disorders, which have been suggested to be divergent, closer together. Moreover, the results of this study could lead to early detection and early intervention for patients with psychiatric disorders. As the concept of duration of untreated psychosis (DUP) suggests [14], early detection and intervention are important for a favorable disease prognosis.

There has been an increase in recent years in the number of reported studies that have attempted to diagnose psychiatric disorders using natural language processing. Target disorders include schizophrenia [15, 16], depression [17], bipolar affective disorder, obsessive-compulsive disorder [18], autism spectrum disorders [19], dementia [20, 21] and many others. Although there are reports of the successful prediction of disease onset with a high accuracy [22], challenges remain in terms of larger sample sizes, the extraction of multiple features that are simple and robust for clinical application, the search for features that are useful across diseases, and investigations of the relationship between the disease time course and disease severity. The present study aims to address these issues. Particularly, this study aims to target 300 to 500 participants and to obtain 600 to 2500 datasets; as such, it will be one of the largest data sets among studies examining the diagnosis of major psychiatric disorders using natural language processing. In this study, we will explore methods that can be used to discriminate between healthy-disease and disease-disease in a large sample size to challenge such issues.

Furthermore, we plan to apply the findings of this research to text data collected from social networking services, such as Twitter. We expect that this would enable us to analyze larger-scale data longitudinally and to infer the mood of society and its relationships among social events. For this purpose, we will also construct our own Japanese sentiment polarity dictionary to conduct a detailed analysis.

The challenges of this research are as follows. First, it will be very important to ensure a similar annotation protocol among the examiners. Similarly, it will be very important to keep the inter-rater reliability high when diagnosing and/or assessing patients. Anticipating this issue, the study team has developed educational modules to maintain a high quality of ratings, and the inter-rater reliability will be tested using a random sampling during the study period. Second, as far as we know this is the first study of this kind to be conducted on such a large sample size in a Japanese-speaking region. To date, most of the reported cases of natural language processing applied to psychiatric disorders have been conducted in English-speaking regions; therefore, a replication study in the same language would be difficult to perform. Some features cannot exist in Japanese due to differences in grammar. For example, the determiners used to analyze linguistic features of psychosis[22] do not exist in Japanese. Moreover, the Japanese language presents a few challenges. First of all, the quantity of publicly available conversational data in the Japanese language is relatively small, and almost none is from speakers with mental disorders. Secondly, the Japanese language is an agglutinative language, in that it does not use spaces between words, so its word boundaries are not as clear as they are in other languages. Another challenge involves handling spoken conversational data, which tends to be broken or ill-formed. Furthermore, the Japanese language has a characteristic of often omitting arguments such as subjects. At the same time, however, if common features capable of identifying specific psychiatric disorders across languages can be found, such information would likely become an important disease feature. Although there are many challenges, this research will enable us to study the languages of psychiatric disorders statistically and comprehensively, and it will provide a new interpretation of language to psychiatrists.

## Data Availability

Data not available due to ethical restrictions

## Ethics approval and consent to participate

This study was approved by the Institutional Review Board of Keio University School of Medicine and the participating medical facilities. Researchers will obtain written informed consent from all the participants. In cases where the patients are judged to be decisionally impaired, the patients’ guardians will be asked to provide consent. Participants will be able to leave the study at any time.

## Preprint and previous presentation

The design of the study and the dataset that was acquired were briefly presented at the International Workshop on Health Intelligence, 2019 [23].

## Funding

This research is supported by the Japan Science and Technology Agency CREST under Grant Number JPMJCR1684 and JPMJCR19F4. A former grant was awarded in 2016 and ended in 2018. A latter grant was awarded in 2019 and will end in 2021. The funding source did not participate in the design of this study and will not be involved in the study’s execution, analyses, or submission of results.

Japan Science and Technology Agency (JST) Kawaguchi Center Bldg. 4-1-8 Honcho, Kawaguchi-shi, Saitama 332-0012 Japan. Tel: +81-48-226-5601, Fax: +81-48-226-5651.

## Authors’ contributions

All the authors participated in the design of the study. TK, HN, YK, KCL, and TY wrote the manuscript, and the remaining authors critically reviewed the manuscript.

## Acknowledgements

We gratefully acknowledge the UNDERPIN collaborators:

Yuki Tazawa, Yuki Ito, Yuriko Kaise, Sayaka Hanashiro, Yoshitaka Yamaoka, Noriko Maegaichi, Kaori Okubo, Kiko Shiga, Sakura Takeuchi, Shimpei Isa, Kelley Cortright (Keio University), Akiko Goto (Tsurugaoka Garden Hospital), Yoshino Humihiro (Tsurugaoka Garden Hospital), Nobuya Ishida (Biwako Hospital), Yuka Oba (Sato Hospital),

## Abbreviations

UNDERPIN: Understanding Psychiatric Illness Through Natural Language Processing
DSM-5: Diagnostic and Statistical Manual of Mental Disorders, Fifth Edition
ICD-11: International Classification of Diseases 11th Revision
UMIN: University Hospital Medical Information Network
M.I.N.I.: Mini-International Neuropsychiatric Interview
MMSE: Mini-Mental State Examination Mini-International; Neuropsychiatric Interview
QOL: Quality Of Life
YLDs: years lived with disability
CGI-S: Clinical General Impression Scale-Severity
SCID: Structural Clinical Interview for DSM-5
HAM-D: Hamilton Depression Rating Scale
MADRS: Montgomery Asberg Depression Rating Scale
YMRS: Young Mania Rating Scale
STAI: State-Trait Anxiety Inventory
PANSS: Positive And Negative Syndrome Scale
MMSE: Mini-Mental State Examination
MoCA-J: Montreal Cognitive Assessment-Japanese version
WMS-RLM: Wechsler Memory Scale-Revised Logical Memory
CDR: Clinical Dementia Rating
F0: fundamental Frequency
F1: 1^st^ formant Frequency
F2: 2^nd^ formant Frequency
F3: 3^rd^ formant Frequency
CPP: Cepstral Peak Prominence
MFCC: Mel-Frequency Cepstrum Coefficients
SVM: support vector machines
DUP: duration of untreated psychosis

